# Social distancing to slow the U.S. COVID-19 epidemic: longitudinal pretest-posttest comparison group study

**DOI:** 10.1101/2020.04.03.20052373

**Authors:** Mark J. Siedner, Guy Harling, Zahra Reynolds, Rebecca F. Gilbert, Sebastien Haneuse, Atheendar S. Venkataramani, Alexander C. Tsai

## Abstract

**Background:** Social distancing measures to address the U.S. coronavirus disease 2019 (COVID-19) epidemic may have notable health and social impacts.

**Methods and Findings:** We conducted a longitudinal pretest-posttest comparison group study to estimate the change in COVID-19 case growth before versus after implementation of statewide social distancing measures in the U.S. The primary exposure was time before (14 days prior to, and up to 3 days after) versus after (beginning 4 days after, and up to 21 days after) implementation of the first statewide social distancing measures. Statewide restrictions on internal movement were examined as a secondary exposure. The primary outcome was the COVID-19 case growth rate. The secondary outcome was the COVID-19-attributed mortality growth rate. All states initiated social distancing measures between March 10-25, 2020. The mean daily COVID-19 case growth rate decreased beginning four days after implementation of the first statewide social distancing measures, by 0.9% per day (95% confidence interval [CI], −1.3% to −0.4%; P<0.001). We did not estimate a statistically significant difference in the mean daily case growth rate before versus after implementation of statewide restrictions on internal movement (0.1% per day; 95% CI, −0.04% to 0.3%, P=0.14), but there is significant difficulty in disentangling the unique associations with statewide restrictions on internal movement from the unique associations with the first social distancing measures. Beginning seven days after social distancing, the COVID-19-attributed mortality growth rate decreased by 1.7% per day (95% CI, −3.0% to −0.7%; P<0.001). Our analysis is susceptible to potential bias resulting from the aggregate nature of the ecological data, potential confounding by contemporaneous changes (e.g., increases in testing), and potential underestimation of social distancing due to spillovers across neighboring states.

**Conclusions:** Statewide social distancing measures were associated with a decrease in the COVID-19 epidemic case growth rate that was statistically significant and a decrease in the COVID-19-attributed mortality growth rate that was not statistically significant.

**Author Summary:** *Why was the study done:* There are few empirical data about the population health benefits of imposing statewide social distancing measures to reduce transmission of severe acute respiratory syndrome coronavirus 2, which causes coronavirus disease 2019 (COVID-19).

*What did the researchers find:* We compared data from each state before vs. after implementation of statewide social distancing measures to estimate changes in mean COVID-19 daily case growth rates. Growth rates declined by approximately 1% per day beginning four days (approximately one incubation period) after statewide social distancing measures were implemented. Stated differently, our model implies that social distancing reduced the total number of COVID-19 cases by approximately 1,600 reported cases at 7 days after implementation, by approximately reported 55,000 cases at 14 days after implementation, and by approximately reported 600,000 cases at 21 days after implementation.

*What do these findings mean:* Statewide social distancing measures were associated with a reduction in the growth rate of COVID-19 cases in the U.S. However, our analysis is susceptible to potential bias resulting from the aggregate nature of the data, potential confounding by other changes that occurred during the study period (e.g., increases in testing), and potential underestimation of social distancing due to spillovers across neighboring states.

## Introduction

Modeling estimates suggest that up to 80% of Americans will be infected with severe acute respiratory syndrome coronavirus 2 (SARS CoV-2) if no preventive interventions are implemented [1]. In response, U.S. state governments have applied social distancing measures in an attempt to limit its transmission and reduce morbidity and mortality from coronavirus disease 2019 (COVID-19). Such measures have been applied during prior pandemics, with moderate success [2-5], and are predicted to prevent a rapid, overwhelming epidemic in modeling studies [6]. Data on social distancing and its associations with the course of the COVID-19 pandemic are now beginning to emerge, although no studies have examined changes in COVID-19-attributed mortality as an outcome. Recent studies have shown slowed COVID-19 epidemic growth coinciding with reductions in internal movement [7,8], as well as after implementation of shelter-in-place orders [9-11]. Because of the economic and social costs associated with social distancing measures [12,13], there is immense value in using different modalities to quantify the extent to which they have benefits for epidemic control. Herein, we contribute to this body of evidence, using a longitudinal pretest-posttest comparison group study design to compare the daily growth rate of COVID-19 cases, and the daily growth rate of COVID-19-attributed deaths, before and after implementation of social distancing measures in the U.S. Our primary aim was to empirically estimate the public health impact of government mandated non-pharmacological interventions in the period after their initial implementation and prior to their recently staged relaxation.

## Methods

### Data Collection

We searched government websites and third-party sources to identify all statewide social distancing measures implemented between January 21 and May 1, 2020 (see **Supplement Table 1**). We applied a broad definition of social distancing measures, using a previously published typology [14]: closures of schools, closures of workplaces, cancellations of public events, restrictions on internal movement, and closures of state borders. Restrictions on internal movement, i.e., shelter-in-place orders (often referred to colloquially as “lockdowns”), are generally the most restrictive of these in terms of their impacts on daily movement. To obtain daily state-specific reported COVID-19 cases and deaths, we used the *New York Times* COVID-19 database (https://github.com/nytimes/covid-19-data, last accessed May 26, 2020). Reporting of cases and deaths in the *New York Times* database varies by state, but typically includes both laboratory-confirmed and suspected cases, as recommended by the Council of State and Territorial Epidemiologists [15].

Our primary outcome was the rate of change in daily COVID-19 cases in each state, calculated as the natural log of cases on each date minus the natural log of cases on the prior date. Analysis was restricted to days in which a state had at least 30 cumulative cases reported, to minimize any effects of volatile rate changes early in the epidemic. The primary exposure of interest was time, measured as a continuous variable and divided into two periods: pre-implementation (14 days prior to, through three days after, implementation of the first statewide social distancing measure) versus post-implementation (beginning 4 days after, and up to 21 days, after implementation). We selected this transition point based on previously published estimates of the lower end of the 95% confidence interval of the COVID-19 incubation period [16-18], which is when cases at a population level should be expected to decline in the setting of a structural intervention. The latent period (between infection and the onset of infectiousness) is shorter than the incubation period (between infection and the onset of symptoms), but during the time period covered by this study -- and as of the time of this writing -- active asymptomatic screening programs have not been implemented in the U.S., so case counts would not have been expected to change until individuals became symptomatic and presented for testing. We limited our analysis to 21 days after implementation to prevent any potential diluting effects resulting from relaxation of some social distancing measures, which in some states began less than four weeks after implementation (e.g.. Alabama, Alaska, Mississippi, and South Carolina).

The secondary outcome was the rate of change in daily COVID-19-attributed deaths in each state, calculated as the natural log of deaths on each date minus the natural log of deaths on the prior date. Analysis was restricted to days in which a state had at least 30 cumulative deaths reported. In the analysis of COVID-19-attributed deaths, there was less certainty about the hypothesized lag between implementation of social distancing and observed changes in daily COVID-19-attributed deaths. The median time from symptom onset to death varies widely in the literature. The COVID-19 Surveillance Group [19] has estimated a median of 8 days from symptom onset to death in Italy, while estimates from China are approximately twice that amount [20-22]. After incorporating a 3-5 incubation period, we hypothesized that a beneficial impact on COVID-19-attributed mortality, if any, would be observed no sooner than 7 days and no later than 14 days after implementation. Therefore, we took a more exploratory approach to modeling the time from enactment of statewide social distancing, selecting time periods for the spline terms (described below) every 3 days during days 4-17 (i.e., 4, 7, 10, 14, and 17 days) and allowed for observation in the post-intervention period up through 30 days.

As a secondary exposure, we examined the dates of statewide restrictions on internal movement. For states that did not implement statewide restrictions on internal movement during the study period, we set day 0 as 14 days after the last state implemented a restriction on internal movement (April 21, 2020) and attributed the prior 14 days to the pre-implementation period.

### Statistical Analysis

We fit mixed effects linear regression models, specifying the log difference in daily cases as the outcome of interest and a random effect for state to allow for within-state correlation of cases over time. Explanatory variables included time in days, implementation period, and a time-by-implementation period product term. This analysis was not conducted as part of a funded study or pre-planned/registered study protocol. However, we followed a clear analysis plan (see **Supplement**), with minor changes made in response to a rapidly changing epidemic and in response to editorial and reviewer feedback. The initial analysis, described in the manuscript we deposited with the *medRxiv* preprint server on April 8, 2020 [23], included data on statewide social distancing measures implemented between January 21 and March 30, 2020, and COVID-19 cases through March 31. Prior to submission of the current manuscript, we updated the dataset and extended the study period to include social distancing measures implemented up to April 8 and COVID-19 cases and deaths up to April 8. In response to editorial and reviewer feedback, we further updated the dataset and extended the study period to include social distancing measures implemented up to May 1 and COVID-19 cases and deaths up to May 26, expanded the analysis to include growth in COVID-19-attributed deaths as a secondary outcome, and added sensitivity analyses investigating a range of plausible incubation periods and an event study specification (see below).

To assess the robustness of our findings, we conducted several sensitivity analyses. First, to adjust for potential confounding by population density, we adjusted our estimates by state-level population density [24,25]. To account for weekly periodicity that could also coincide with implementation of social distancing measures, we also adjusted for day of the week [26,27]. Second, to assess the extent to which early vs. late implementation of social distancing measures modified the effects of these actions [28], we stratified our estimates by the size of the epidemic in the state. Third, although implementation of social distancing measures is most likely to show population-level effectiveness in reducing coronavirus transmission beginning 3 days later at the lower bound of its estimated incubation period [16], we refitted the regression models specifying a range of different incubation periods.

Finally, to assess the extent to which our estimates may have been driven by model specification, we replaced the longitudinal pretest-posttest comparison group study design with a multivariable regression model in which the primary explanatory variables of interest were a series of binary indicators denoting each day before versus after implementation of the first statewide social distancing measures (often described in the econometrics literature as an event study specification [29-31]). This approach compares daily case growth before versus after implementation of the first statewide social distancing measures in states that implemented such measures versus daily case growth in states that did not implement such measures. Our regression model included state fixed effects, to adjust for potential confounding from time-invariant state-level factors or baseline differences in population socioeconomic or health characteristics; and linear and quadratic terms for time (days) to adjust for nationwide secular trends in the outcomes. We computed 95% confidence intervals adjusted for clustering within states, the geographical level at which exposure occurred [32].

### Ethics Statement

This ecological analysis was based on publicly available data and was exempt from ethical review.

## Results

A complete list of dates of statewide social distancing measures, by type of measure and state, is contained in the **Supplement** (**Supplement Table 1**). During March 10–25, all 50 states and the District of Columbia implemented at least one statewide social distancing measure (**Supplement Figure 1**). The most widely enacted measures on the first date of implementation were cancellations of public events (34/51 [67%]) and closures of schools (26/51 [51%]). The first social distancing measures were implemented when the median epidemic size was 35 cases (interquartile range [IQR], 17-72).

**Figure 1A** shows the mean COVID-19 daily case growth rate mapped against the date of the first statewide distancing measures. At the date of implementation of the first social distancing measure, states had a mean daily case growth rate of 30.8% (95% confidence interval [CI], 29.1-32.6; **Table 1**), corresponding to a doubling of total cases every 3.3 days. From fourteen days prior, through three days after, implementation of the first social distancing measure, the mean daily case growth rate did not change (-0.2% per day; 95% CI, −0.6% to 0.3%; *P*=0.51). Beginning four days after implementation of the first statewide social distancing measure, the mean daily case growth rate decreased by an additional 0.9% per day (95% CI, −0.4% to −1.4%; *P*<0.001). This estimate corresponds to a mean daily case growth rate that had declined to 26.5% (doubling of total cases every 3.8 days) by day 7 after enactment of the first statewide social distancing measures, to 19.6% (doubling time of 5.1 days) by day 14, and to 12.7% (doubling time of 7.9 days) by day 21.

**Figure 1A-B.**
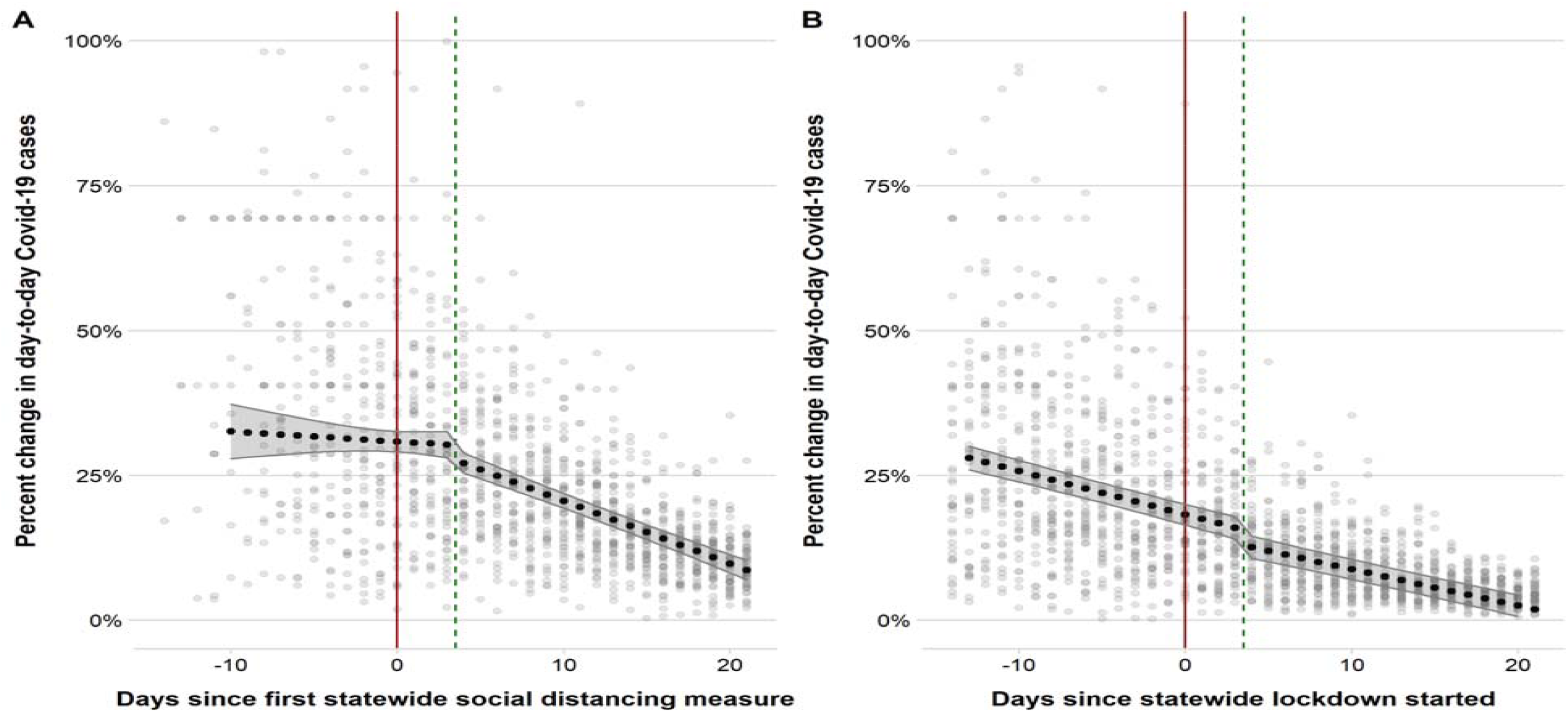
Scatter plots and predictive margins with 95% confidence interval derived from regression models of the daily COVID-19 growth rate pre vs. post-implementation of the first statewide social distancing measures (1A) and statewide restrictions on internal movement (1B). The red line indicates the date of implementation in each state. The green dashed line is 4 days after implementation of the social distancing measure.

**Table 1.** Linear regression models for the growth in mean daily COVID-19 cases before vs. after implementation of the first statewide social distancing measure and statewide restrictions on internal movement

|  | First statewide social distancing measure |  |  | Statewide restriction on internal movement |  |  |
| --- | --- | --- | --- | --- | --- | --- |
|  | **b** | **95% CI** | **P-value** | **b** | **95% CI** | **P-value** |
| Constant | 0.309 | 0.291, 0.326 | <0.001 | 0.183 | 0.164, 0.201 | <0.001 |
| Time (days relative to intervention) | -0.002 | -0.006, 0.003 | 0.44 | -0.008 | -0.009, -0.006 | <0.001 |
| Post-intervention period | 0.006 | -0.017, 0.029 | 0.59 | -0.031 | -0.047, -0.015 | <0.001 |
| Time × post-intervention period | -0.009 | -0.013, -0.004 | <0.001 | 0.001 | -0.00, 0.003 | 0.14 |

*Notes: b, estimated regression coefficient. CI, confidence interval. The post-intervention period begins four days after social distancing measures were implemented to account for the disease incubation period.*

As of May 1, nearly all (45 [90%]) states had implemented a statewide restriction on internal movement. These restrictions on internal movement were implemented a median of 11 days after the first statewide social distancing measure (IQR 8-15) was implemented in the respective states, when the median epidemic size was 937 cases (IQR, 225-1414). The mean daily case growth rate had already been declining at a mean rate of −0.8% per day during the 14 days prior to implementation of statewide restrictions on internal movement (95% CI, −0.9% to −0.7%; *P*<0.001) (**Table 1, Figure 1B**). There was a drop detected three days after statewide restrictions on internal movement were implemented (-3.1%; 95% CI, −4.7% to −1.5%, *P*<0.001), but no statistically significant difference in the rate of change from before versus after their implementation (0.1% per day; 95% CI, −0.04% to 0.3%, *P*=0.14). As discussed in more detail below, there is substantial difficulty in disentangling the unique associations with statewide restrictions on internal movement from the unique associations with the first social distancing measures.

In the analysis of the secondary outcome, change in daily COVID-19-attributed deaths, given the uncertainty in the hypothesized lag between implementation of social distancing and observed changes (if any) in daily COVID-19-attributed deaths, we explored a range of thresholds. As shown in **Table 2**, by seven days after implementation of the first statewide social distancing measure, the mean daily growth rate in COVID-19-attributed deaths decreased by 2.0% per day (95% CI, −3.0% to −0.9%; *P*<0.001). By 14 days the estimated association was no longer statistically significant (-1.0% per day; 95% CI, −0.2% to 0.1%; *P*=0.09). No additional statistically significant benefit was found after 7 days of implementation of statewide restrictions on internal movement.

**Table 2.**
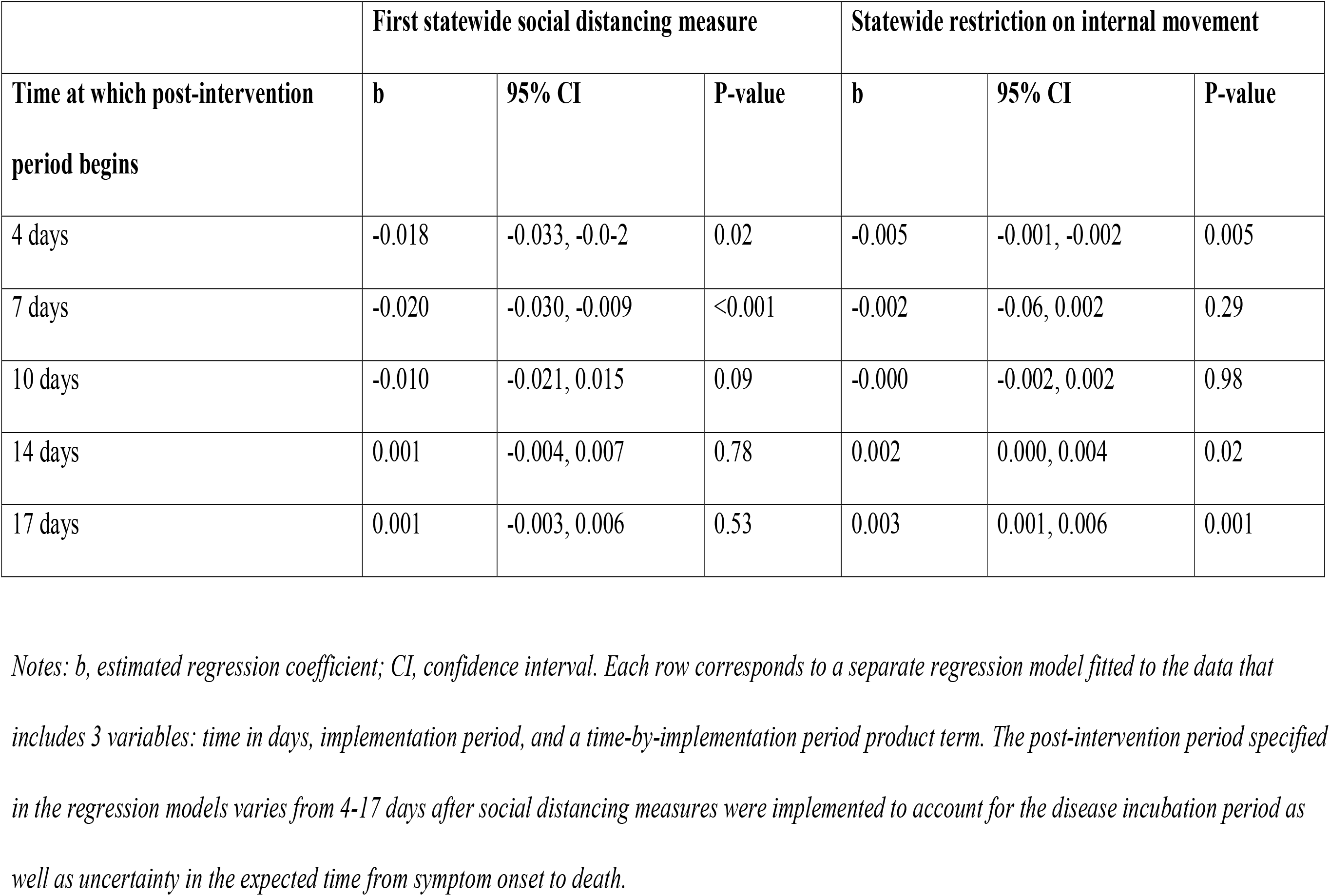
Linear regression models for growth in mean daily COVID-19-attributed deaths before vs. after implementation of the first statewide social distancing measure and statewide restrictions on internal movement, assuming a range of days between symptom onset and death

|  | First statewide social distancing measure |  |  | Statewide restriction on internal movement |  |  |
| --- | --- | --- | --- | --- | --- | --- |
| Time at which post-intervention period begins | b | 95% CI | P-value | b | 95% CI | P-value |
| 4 days | -0.018 | -0.033, -0.0-2 | 0.02 | -0.005 | -0.001, -0.002 | 0.005 |
| 7 days | -0.020 | -0.030, -0.009 | <0.001 | -0.002 | -0.06, 0.002 | 0.29 |
| 10 days | -0.010 | -0.021, 0.015 | 0.09 | -0.000 | -0.002, 0.002 | 0.98 |
| 14 days | 0.001 | -0.004, 0.007 | 0.78 | 0.002 | 0.000, 0.004 | 0.02 |
| 17 days | 0.001 | -0.003, 0.006 | 0.53 | 0.003 | 0.001, 0.006 | 0.001 |

*Notes: b, estimated regression coefficient; CI, confidence interval. Each row corresponds to a separate regression model fitted to the data that includes 3 variables: time in days, implementation period, and a time-by-implementation period product term. The post-intervention period specified in the regression models varies from 4-17 days after social distancing measures were implemented to account for the disease incubation period as well as uncertainty in the expected time from symptom onset to death.*

Sensitivity analyses suggested our estimates were not sensitive to inclusion of additional covariates, did not differ by size of the epidemic at implementation, and were consistent with the known incubation period (**Supplement Tables 2, 3**, and **4**). In the event study specification (**Supplement Figure 2**), mean daily case growth was negative by day 4, and the estimates were statistically significant by day 8, consistent with the primary analysis. The event study analysis for change in daily COVID-19-attributed deaths also produced estimates qualitatively similar to the primary analysis, although with slightly larger confidence intervals given the smaller number of events (**Supplement Figure 3**).

## Discussion

In this longitudinal pretest-posttest comparison group study, we found that implementation of social distancing measures was associated with a reduction in the mean daily growth rate of COVID-19 cases and in the mean daily growth rate of COVID-19-attributed deaths. Our estimates imply a more than doubling in the doubling time (from 3.8 days to 8.0 days) by three weeks following the implementation of social distancing measures. Assuming a cumulative epidemic size of 4,125 reported cases (equivalent to the cumulative number of cases in the U.S. at the time of implementation in each state), the reduction in growth rate we estimated corresponds to a difference between 26,281 reported cases with no social distancing versus 24,625 reported cases at 7 days after implementation, a difference between 158,518 reported cases with no social distancing versus 102,223 reported cases at 14 days after implementation, and a difference between 904,773 reported cases with no social distancing versus 283,161 reported cases at 21 days after implementation. Stated differently, our model implies that social distancing reduced the total number of reported COVID-19 cases by approximately 1,600 cases at 7 days after implementation, by approximately 56,000 reported cases at 14 days after implementation, and by approximately 621,000 reported cases at 21 days after implementation.

These results are consistent with both the theoretical effect of social distancing on epidemic spread [6] and the historical benefit observed with the implementation of such interventions during prior epidemics of communicable diseases [28]. They also are largely in keeping with recent data on the impacts of social distancing measures in the U.S. on both reduced mobility [7,8], and case growth rates [9-11], with generally similar effect sizes. Our study extends this literature by further examining COVID-19-attributed mortality as an outcome. The associations between social distancing and case growth rates were most apparent at the lower end of the incubation period that has been estimated based on publicly available data, with some evidence that the change in growth rates may have started even earlier. We suspect that this observation may have resulted from self-imposed social distancing that has reportedly occurred prior to government-issued mandates [33].

Our findings should be interpreted with the following limitations in mind. Our estimates would be biased toward the null if: 1) state and local governments had intensified social distancing measures in response to a worsening epidemic, 2) there were substantial violations of the stable unit treatment value assumption (e.g., workplace closures of large employers that had spillover effects across state lines), or 3) surveillance and testing intensified during the study period (thereby resulting in increased case reporting). Moreover, statewide restrictions on internal movement were often implemented after other social distancing measures had already been applied, further biasing our estimate toward the null. Estimates of cases and deaths in our model include those that are both laboratory-confirmed and suspected by health departments, but they are both likely to be under-estimates, due to limitations in testing, the presence of asymptomatic cases, and the occurrence of deaths that are not attributed to COVID-19 [34,35]. Nonetheless, our analyses focus on day-to-day changes in the growth rate of cases and deaths, so underreporting would only bias our results if reported versus true outcomes systematically differed from prior to versus after the implementation of social distancing measures. In contrast, our projected estimates of cases prevented are likely to be highly conservative, because they are modeled based on reported cases. While some studies have suggested that cases have been under-reported in the U.S. by 1-2 orders of magnitude [36], deaths appear to be under-counted by approximately 25% [34]. Finally, our estimates of the associations between social distancing and changes in mean daily case growth rates (and the corresponding number of cases averted) cannot be extrapolated linearly beyond the 21-day post-implementation analysis window. At the time of this analysis, states had begun to enact social relaxation measures, which necessarily prevents drawing conclusions about the long-term associational effects of these interventions in isolation.

Our study does not ascertain which of the statewide social distancing measures were most effective in reducing mean daily COVID-19 case growth. A variety of social distancing measures were used, often simultaneously, making it difficult to disentangle their independent associations. It is also possible that some state residents changed their behaviors in response to local (e.g., county-level) social distancing measures enacted prior to the statewide measures, or that some state residents changed their behaviors independently of social distancing measures enacted at any level of jurisdiction. The latter phenomenon has been observed empirically in other settings [37,38]. Such behavior change would be consistent with our sensitivity analysis suggesting a signal that extends back to a 0-day incubation period.

Finally, our analysis cannot answer questions about the appropriate time for rescinding social distancing measures. While our findings demonstrate that early mitigation efforts have yielded a substantial population health benefit, these benefits should be weighed against their costs. The costs of social distancing are likely to exacerbate the confluence of longstanding economic, social, and health decline that is already occurring in the U.S. [39-41], brought into even sharper relief given emerging data about racial, ethnic, and socioeconomic disparities in the incidence of COVID-19-related burden [42,43]. Moreover, there is currently robust debate about the extent to which legal authority to initiate or rescind social distancing measures resides with the federal or local government [44]. Additional study may be possible in the coming weeks to months if some statewide social distancing measures are relaxed while others are retained, or if measures are relaxed in some local jurisdictions while being retained in others. The use of cross-country data may also be helpful in this regard, albeit limited by any substantial within-country heterogeneity (such as has been observed in the diversity of uncoordinated U.S. federal and state responses to the epidemic).

Our finding that implementation of statewide social distancing was associated with a reduction in the mean daily growth rate of COVID-19-attributed deaths should be interpreted with more caution, given the uncertainty in published estimates about the median time from symptom onset to death [19-21]. The strongest associations we estimated, in terms of both statistical significance and magnitude, occurred at seven days after implementation. While this change could be reflective of what is known about time to death for the median hospitalized patient, other plausible thresholds were not associated with a statistically significant reduction in the mean daily growth rate of COVID-19-attributed deaths.

The finding that implementation of statewide restrictions on internal movement was associated neither with a statistically significant reduction in the mean daily growth rate of COVID-19 cases nor with a statistically significant reduction in the mean daily growth rate of COVID-19-attributed deaths warrants additional scrutiny. Identification of the unique effect of statewide restrictions on internal movement, separate from the initial social distancing measures, is a difficult undertaking given that they were implemented a median of 10 days later. There were no states in which statewide restrictions on internal movement were implemented without the prior implementation of other social distancing measures (e.g., cancellations of public events and closures of schools). As a result, we could not test hypotheses about the independent causal effects of statewide restrictions on internal movement, but rather could only ascertain the extent to which they were associated with additional reductions in the COVID-19 case growth rate, i.e., beyond those generated by the initial statewide social distancing measures. Moreover, the null finding should be interpreted in light of the other likely biases toward the null previously discussed.

In summary we demonstrate that the U.S. COVID-19 epidemic growth rate began to decline within approximately one incubation period following the initiation of statewide social distancing measures. Exploratory findings suggest that social distancing may also have had lagged mortality benefits. Future work should consider the potential public health benefits of relaxing these measures and, specifically, to what extent and how differential relaxation of certain measures promotes persistent epidemic control.

## Data Availability

All data will be available through public databases and the supplementary materials.

